# Identification on Admission of COVID-19 Patients at Risk of Subsequent Rapid Clinical Deterioration

**DOI:** 10.1101/2020.08.13.20171751

**Authors:** J. Beals, J. Barnes, D. Durand, J. Rimar, T. Donohue, M. Hoq, K. Belk, A. Amin, M.J. Rothman

## Abstract

**Introduction:** Recent localized surges in COVID-19 cases have resulted in the hospitals serving those areas being overwhelmed. In such cases, the ability to rapidly and objectively determine a patient’s acuity and predict near-term care needs is a major challenge. At issue is the clinician’s ability to correctly identify patients at risk for subsequent rapid clinical deterioration. Data-driven tools that can support such determinations in real-time may be a valuable adjunct to clinician judgement during COVID-19 surges.

**Objective:** To assess the effectiveness of the Rothman Index (RI) predictive model in distinguishing the risk of subsequent deterioration or elevated care needs among hospitalized COVID-19 patients at the time of hospital admission.

**Methods:** We evaluated the initial RI score on admission to predict COVID-19 patient risk for 216 COVID-19 patients discharged from March 21^st^ to June 7^th^, 2020 at Sinai LifeBridge Hospital and 1,453 COVID-19 patients discharged from any of Yale New Haven Health System’s Yale New Haven, Bridgeport, and Greenwich hospitals from April 1^st^ to April 28^th^, 2020. In-hospital mortality as a function of age and RI on admission for COVID-19 and non-COVID-19 patients were compared. AUC values using each COVID-19 patient’s initial RI on admission to predict in-hospital mortality, mechanical ventilation, and ICU utilization were computed, as were precision and recall for mortality prediction at specific RI thresholds.

**Results:** The RI computed at the time of admission provides a high degree of objective discrimination to differentiate the COVID-19 population into high and low risk populations at the outset of hospitalization. The high risk segment based on initial RI constitutes 20-30% of the COVID-19 positive population with mortality rates from 40-50%. The low risk segment based on initial RI constitutes 40%-55% of the population with mortality rates ranging from 1%-8%. Of note is that COVID-19 patients who present with heightened but generally unremarkable acuity can be identified early as having considerably elevated risk for subsequent physiological deterioration.

**Conclusion:** COVID-19 patients exhibit elevated mortality rates compared to non-COVID-19 medical service patients and may be subject to rapid deterioration following hospital admission. A lack of predictive indicators for identifying patients at high risk of subsequent deterioration or death can pose a challenge to clinicians. The RI has excellent performance characteristics when stratifying risk among COVID-19 patients at the time of admission. The RI can assist clinicians in real-time with a high degree of objective discrimination by segmenting the COVID-19 population into high and low risk populations. This supports rapid and optimal patient bed assignment and resource allocation.

## Introduction

The risk to patients posed by the COVID-19 disease (caused by the severe acute respiratory syndrome coronavirus 2 SARS-CoV-2) varies greatly among individuals. Somewhere between 5-20% of those infected become critically ill with advanced age, chronic diseases and co-morbidities being substantial risk factors.^1^ Hospitalized COVID-19 patients are susceptible to rapid deterioration and onset of acute respiratory distress syndrome which may require respiratory support and endo-tracheal intubation.^2,3^ These patients exhibit a high rate of intensive care unit (ICU) utilization, mechanical ventilation, and significant risk of in-hospital mortality.^4^

In regions which have seen significant COVID-19 case-rates in the general population, as in the Lombardy region of Italy or the New York City region of the United States, local hospitals have been overwhelmed by the surge in patient volumes.^5-7^ These surge conditions can result in fatigued, over-extended clinical staff having to make rapid and accurate assessment of patient risk – a difficult task during the best of times – in order to determine the allocation of limited hospital resources, e.g. mechanical ventilators and ICU beds. The professional and personal challenges faced by caregivers having to allocate scarce resources under pandemic conditions cannot be overstated.^8^ Use of data-driven, validated models to support the triaging of COVID-19 patients has the potential to ease the burden on clinical staff by supporting them in making difficult care decisions in a more effective and objective fashion.

Current reports indicate that age and comorbidity correlate with mortality risk.^9^ We postulate that both are in essence being treated as indirect estimates of patient acuity. However, age itself is not intrinsically a risk factor but rather a proxy for age-related diseases and infirmities. Co-morbidity is a more explicit measure of diseases and infirmities, but the presence or absence of co-morbidities does not directly translate to acuity or COVID-19 mortality risk without characterization of the current impact of the disease. Our work bypasses the need for a proxy-based approach by using the Rothman Index (RI) to directly measure physiologic acuity.

While numerous expert-based and machine learning derived patient scoring systems exist,^10,11^ few are validated in clinical practice and fewer still have been evaluated for use with COVID-19 patients. To stratify the risk of COVID-19 patients at the time of admission, we evaluate the performance of the RI – an extensively validated patient condition score (PeraHealth, Inc. Charlotte NC) in clinical use by multiple hospitals and health systems.

The RI synthesizes a wide range of discrete clinical inputs including vital signs, summarized head-to-toe nursing assessments, and lab values to calculate an overall acuity score that is updated in real-time as new patient data become available.^12^ These data are routinely documented on all hospitalized patients and deviations from normal values serve as important indicators of deterioration.

The RI is unique in its inclusion of the full range of body-system nursing assessment data – categorized physical and behavioral evaluations of dozens of factors (e.g., patient stops eating, becomes confused, has trouble walking, develops edema, etc.). The deficits charted by nurses are routinely collected on all hospitalized inpatients and serve as important early indicators of deterioration before vital sign derangement or overall clinical worsening. The value of nursing assessment data in predicting adverse outcomes such as mortality has been established,^13^ and their inclusion is one of the reasons that the Rothman Index performs significantly better than acuity scores that are predominantly vital sign based.^14^

In contrast to other commercial early-warning systems, the RI model methodology has been extensively studied and widely published in the literature.^12-16^ It has been validated as an indicator of patient acuity and early deterioration for both pediatric and adult patients across all hospital care settings, e.g., general, intermediate, and intensive care units, and both within and across different patient types and diagnostic groups.^17-20^

With appropriate protocols and workflow integration, the RI’s utility in pro-actively directing clinicians to the bedside has been proven to reduce unplanned transfers to the ICU^21^ and reduce both sepsis associated mortality as well as overall hospital mortality.^22^ In a large unit-randomized controlled trial at Houston Methodist Hospital, a 900-bed academic medical center, implementation of the RI contributed to a 30% reduction in risk-adjusted in-hospital mortality.^23^ Use of the RI for guiding pro-active palliative care consults in ICUs and step-down units can reduce ICU length of stay and associated healthcare utilization and cost.^24^

## Methods

### Data

We analyzed data from Sinai LifeBridge Hospital in Baltimore, MD and from three Yale New Haven Health System (YNHHS) hospitals in New Haven, Bridgeport, and Greenwich CT. These include urban and suburban community hospitals and an academic medical center.

Data from Sinai LifeBridge Hospital included 216 COVID-19 patients discharged between March 21^st^ – June 5^th^, 2020 and 4,108 non-COVID-19 patients discharged between January 2^nd^ – June 7^th^, 2020. Data from the YNHHS hospitals included 1,453 COVID-19 patients discharged between April 1^st^ – April 28^th^, 2020 and 10,093 non-COVID-19 patients admitted and discharged between February 15^th^ – April 28^th^, 2020. At all hospitals, COVID-19 patients were diagnosed on the basis of a lab test and patients identified in the data either by indicators in the patient chart or ICD-10 code U07.1 when available. Starting dates for COVID-19 patient data reflect when such patients were first identified at each hospital.

Inclusion was limited to patients 18 years old and older admitted to a medical service at one of the included hospitals. More than 97% of adult medical patients at each hospital had RI scores. This research was approved by both the Sinai LifeBridge and Yale New Haven Health Bridgeport Institutional Review Boards.

Sinai LifeBridge has the RI software integrated with their Cerner Systems EMR and clinically deployed since 2017. YNHHS has the RI integrated with their Epic Systems EMR and clinically deployed since 2011 (originally with the Allscripts EMR). At all hospitals, RI scores are automatically computed for all patients in all bed locations and automatically updated when new data inputs are documented in the EMR. RI scores were calculated in real-time throughout each patient’s hospitalization and extracted for analysis following the patient’s discharge.

### Characterizing the COVID-19 population

We report the mean and median age, mean and median length of stay, sex and mean Charlson Comorbidity Index (CCI) for COVID-19 and non-COVID-19 patients at all four hospitals. The proportion of COVID-19 and non-COVID-19 patients presenting with a range of common comorbidities as determined using the CCI classification of ICD-10 codes is reported.^25^ To these we added hypertension and obesity based on ICD-10 codes. We additionally computed the CCI for both COVID-19 and non-COVID-19 patients who expired in the hospital to determine any differences between these groups on the basis of comorbidities.

### Quantifying clinician challenges in determining COVID-19 risk

Early research done in China identified patient age as a risk factor for COVID-19 patients and also noted that the initial presentation of COVID-19 was not necessarily indicative of subsequent deterioration or severity of disease progression.^26,27^ The surprising observation regarding initial acuity of hospitalized COVID-19 patients implies a different risk profile than comparable initial acuity for non-COVID-19 patients. To elucidate these differences, we compared the initial RI on admission to patient age on admission, as means for stratifying risk of in-patient mortality for both COVID-19 and non-COVID-19 patients. We further evaluated the initial RI score for both COVID-19 and non-COVID-19 patients against length of stay and ICU utilization. ICU utilization was determined from hospital ADT data, and average ICU hours were calculated based on the total hours of ICU utilization across the complete stay of all patients in the patient population.

### Evaluation of RI for triaging patient risk on admission

RI scores have a maximum value of 100 (no physiologic impairment) with decreasing scores corresponding to increasing acuity. The initial RI score calculated during the admission of each COVID-19 positive patient was evaluated to determine how it performs as an indicator of patient risk.

- *Raw RI Score A UC* – The receiver operating characteristic area under the curve (AUC) was calculated for three independently evaluated end-points: whether the patient (a) expired in the hospital, (b) spent any time in the ICU, (c) required mechanical ventilation.
- *RI vs. Age vs. Comorbidity -* To assess the effectiveness of age, comorbidity and acuity in predicting in-patient mortality, we constructed single-variable logistic regression models for each and compare their performance as measured by AUC.
- *RI-Threshold-based performance -* The precision (positive predictive value) and recall (sensitivity) values at different RI threshold values were evaluated, and two threshold values were chosen, allowing patients to be categorized as being at either low or high risk for subsequent deterioration.

### Opportunity to augment level of care decisions

In order to evaluate whether RI-based risk stratification could provide new insight to providers making care decisions, we analyzed level of care assignment at admission. For patients meeting the RI high risk criteria on admission we evaluated whether patients were admitted directly to ICU, transferred to ICU following admission (and if so how long after admission) or never admitted to an ICU, to determine potential opportunities for improved location assignment or timeliness on the basis of admission risk.

### Evaluation of 24-hour in-patient mortality

To confirm the validity of the RI when applied to COVID-19 patients, we evaluated all RI scores calculated throughout each patient’s hospitalization as a predictor of mortality within 24 hours, and compared the predictive performance of the RI for the COVID-19 population relative to the general medical non-COVID-19 population.

## Results

### Demographics

Population summary characteristics of the COVID-19 patient populations at each hospital are given in Table 1.

**Table 1.**
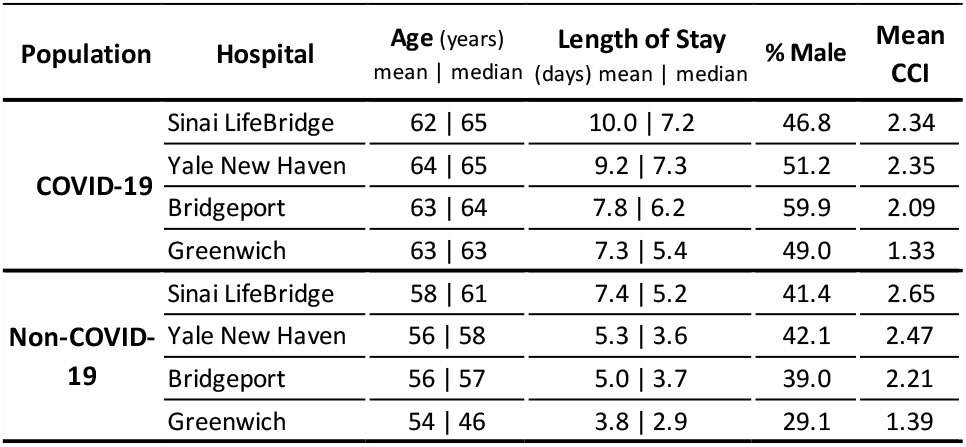
Patient Age, Length of Stay, Sex, CCI.

| Population | Hospital | Age (years) |  | Length of Stay (days) |  | % Male | Mean CCI |
| --- | --- | --- | --- | --- | --- | --- | --- |
|  |  | mean \| median |  | mean \| median |  |  |  |
| COVID-19 | Sinai LifeBridge | 62 \| 65 |  | 10.0 \| 7.2 |  | 46.8 | 2.34 |
|  | Yale New Haven | 64 \| 65 |  | 9.2 \| 7.3 |  | 51.2 | 2.35 |
|  | Bridgeport | 63 \| 64 |  | 7.8 \| 6.2 |  | 59.9 | 2.09 |
|  | Greenwich | 63 \| 63 |  | 7.3 \| 5.4 |  | 49.0 | 1.33 |
| Non-COVID-19 | Sinai LifeBridge | 58 \| 61 |  | 7.4 \| 5.2 |  | 41.4 | 2.65 |
|  | Yale New Haven | 56 \| 58 |  | 5.3 \| 3.6 |  | 42.1 | 2.47 |
|  | Bridgeport | 56 \| 57 |  | 5.0 \| 3.7 |  | 39.0 | 2.21 |
|  | Greenwich | 54 \| 46 |  | 3.8 \| 2.9 |  | 29.1 | 1.39 |

The COVID-19 population is older, more likely to be male and has a longer length of stay. Of note is that the mean CCI is higher across the non-COVID-19 population than for the COVID-19 population.

### Comorbidities

Table 2 shows that preponderant comorbidities include COPD, diabetes, hypertension, and renal disease, in agreement with other reports.^3,9,28^

**Table 2.** Proportion of non-COVID-19 and COVID-19 patients with major or common comorbidities.

| Hospital: | Sinai LifeBridge |  | Yale New Haven |  | Bridgeport |  | Greenwich |  |
| --- | --- | --- | --- | --- | --- | --- | --- | --- |
| Population: | non-COVID-19 | COVID-19 | non-COVID-19 | COVID-19 | non-COVID-19 | COVID-19 | non-COVID-19 | COVID-19 |
| Patient Count: | 4,108 | 216 | 6,757 | 696 | 2,070 | 428 | 1,266 | 329 |
| AIDS/HIV | 2.4% | 0.9% | 0.4% | 1.0% | 0.4% | 0.5% | 0.0% | 0.3% |
| Cancer | 7.1% | 3.2% | 11.6% | 4.3% | 6.8% | 2.1% | 6.0% | 0.9% |
| Cerebrovascular Disease | 13.9% | 8.3% | 7.1% | 4.0% | 5.4% | 4.2% | 4.1% | 2.1% |
| Chronic Pulmonary Disease | 26.5% | 21.8% | 30.2% | 26.3% | 28.7% | 23.6% | 17.1% | 18.2% |
| Congestive Heart Failure | 22.5% | 16.7% | 20.9% | 20.5% | 22.3% | 17.5% | 14.6% | 9.1% |
| Dementia | 7.8% | 22.7% | 6.3% | 17.4% | 10.5% | 19.6% | 8.7% | 12.2% |
| Diabetes with Complications | 15.9% | 19.0% | 13.9% | 17.5% | 15.3% | 16.8% | 7.1% | 9.1% |
| Diabetes without Complications | 22.5% | 35.6% | 17.8% | 29.6% | 18.5% | 27.1% | 8.4% | 22.5% |
| Hypertension | 34.4% | 45.8% | 29.2% | 39.9% | 26.5% | 39.5% | 19.7% | 36.2% |
| Metastatic Carcinoma | 3.8% | 0.5% | 5.6% | 2.2% | 2.5% | 0.9% | 2.3% | 0.6% |
| Mild Liver Disease | 6.7% | 4.2% | 7.7% | 4.9% | 6.3% | 3.7% | 3.4% | 1.2% |
| Moderate or Severe Liver Disease | 1.9% | 1.4% | 2.6% | 0.9% | 2.2% | 0.7% | 1.2% | 0.3% |
| Myocardial Infarction | 11.4% | 11.1% | 8.8% | 10.8% | 8.5% | 5.6% | 5.2% | 6.7% |
| Obesity | 15.4% | 22.2% | 14.3% | 27.2% | 16.7% | 27.1% | 9.6% | 15.8% |
| Paraplegia and Hemiplegia | 5.0% | 1.9% | 2.3% | 1.1% | 1.9% | 1.6% | 1.0% | 1.2% |
| Peptic Ulcer Disease | 1.4% | 1.9% | 1.2% | 1.0% | 1.1% | 0.2% | 0.5% | 1.5% |
| Peripheral Vascular Disease | 10.3% | 6.0% | 7.7% | 5.3% | 7.3% | 6.3% | 3.6% | 3.3% |
| Renal Disease | 20.1% | 21.8% | 18.0% | 21.8% | 18.6% | 22.9% | 12.2% | 12.8% |
| Rheumatic Disease | 2.6% | 1.4% | 3.5% | 4.2% | 3.2% | 3.3% | 3.1% | 2.1% |

In terms of relative occurrence of comorbidities, dementia, diabetes, obesity and hypertension are proportionally over-represented in the COVID-19 population at all four sites.

We computed the mean CCI for both COVID-19 and non-COVID-19 patients who expired in the hospital (Table 3).

**Table 3.** Mean Charlson Comorbidity Index for expired COVID-19 and non-COVID-19 patients.

| Hospital | COVID-19 expired | non-COVID-19 expired |
| --- | --- | --- |
| Sinai LifeBridge | 3.5 | 4.9 |
| Yale New Haven | 3.8 | 4.9 |
| Bridgeport | 3.6 | 4.4 |
| Greenwich | 2.6 | 3.8 |

As with the total COVID-19 and non-COVID-19 populations we see that even focusing on just those patients who expired in the hospital, non-COVID-19 patients have a higher mean CCI than COVID-19 patients.

### Quantifying clinician challenges in determining COVID-19 risk

Patient age has been identified as a risk factor for COVID-19 patients. We evaluated inhospital mortality as a function of initial RI on admission and age as shown in Figure 1.

**Figure 1.**
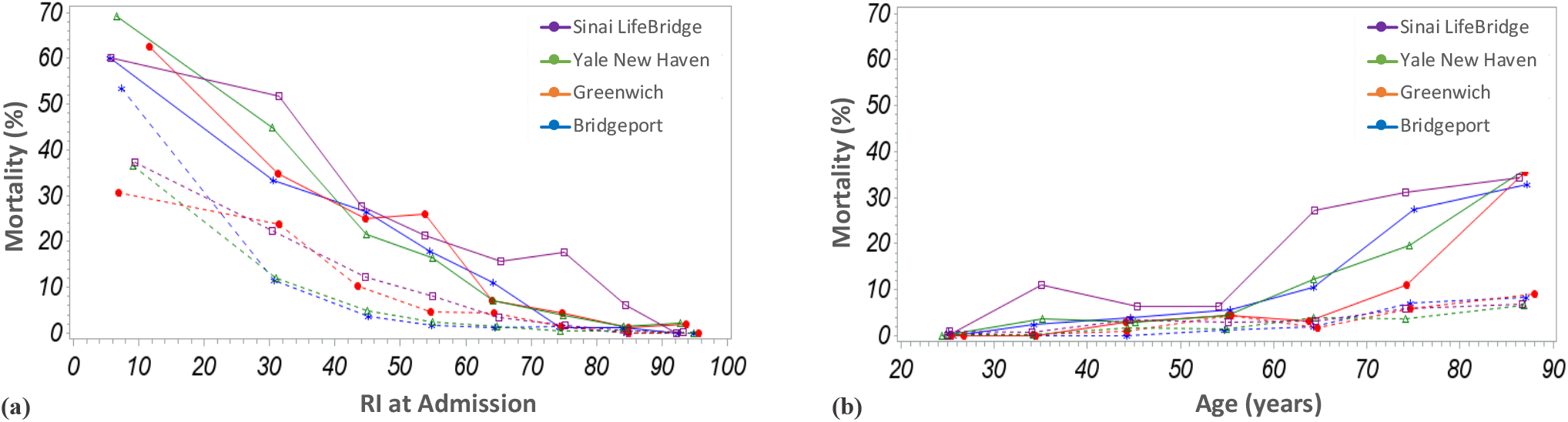
For COVID-19 (solid lines) and non-COVID-19 (dashed lines) patients (a) Mortality vs RI at admission, (b) Mortality vs Age at admission.

The greater range of mortality values on the RI plot relative to the age plot suggests that the RI on admission is more effective than age at separating mortality risk for both COVID-19 and non-COVID-19 patients. Additionally, for each hospital, the separation of the lines between the COVID-19 and non-COVID-19 populations is more distinct over the range of RI values – Figure 1(a), than over the range of age values – Figure 1(b), showing that the RI allows a better discrimination than age of the risk of subsequent deterioration and death.

Figure 2 shows that despite considerable variability in ICU utilization, for COVID-19 and non-COVID-19 patients with similar levels of acuity on admission as measured by the RI, at each hospital COVID-19 patients have comparable or higher degrees of ICU utilization as well as longer lengths of stay. The downturn in both ICU utilization and LOS for lower initial RI scores (below an RI of around 30), corresponds to elevated mortality risk at these acuities, with higher in-hospital mortality resulting in a decrease in the duration of ICU utilization and the hospital stay overall.

**Figure 2.**
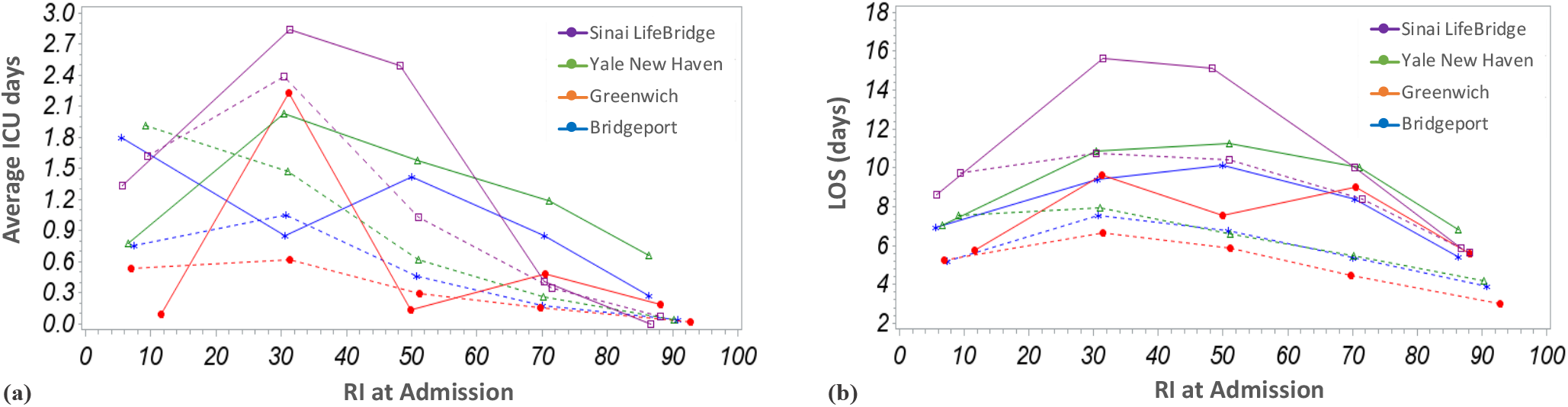
For COVID-19 (solid lines) and non-COVID-19 (dashed lines) patients (a) Average ICU days per patient vs RI at admission, (b) LOS vs RI at admission.

It is interesting to note the convergence of ICU utilization and length of stay for COVID-19 and non-COVID-19 patients at the acuity extrema. That is, COVID-19 and non-COVID-19 patients who are either relatively well or extremely sick at the time of admission have comparable ICU and length of stay outcomes. This supports anecdotal evidence that front-line clinicians don’t tend to struggle with identification of relatively well and extremely sick COVID-19 patients, but are challenged in accurately discerning risk for patients who fall somewhere in the middle, where the apparent acuity of COVID-19 patients does not correspond to typical clinical expectations of patient risk.^29^

### Evaluation of RI for triaging patient risk on admission

To analyze the RI on admission for applicability in admission triage, we used three acuity related endpoints: in-hospital mortality, ICU admission during hospitalization, and mechanical ventilation. The characteristics of the populations with these three outcomes are shown in Table 4 (hospitals are re-ordered and anonymized to blind hospital-level outcomes).

**Table 4.** Percentage of COVID-19 and non-COVID-19 populations expiring, requiring ICU level care, or mechanical ventilation.

| Population | Hospital | Total Expired | ICU Utilization | with ICU who Expired | Mechanical Ventilation | with Ventilation who Expired |
| --- | --- | --- | --- | --- | --- | --- |
| COVID-19 | Hospital A | 15.6% | 26.9% | 23.5% | 14.2% | 37.7% |
|  | Hospital B | 16.3% | 25.9% | 32.8% | 10.8% | 53.3% |
|  | Hospital C | 22.2% | 28.7% | 62.9% | 21.3% | 76.0% |
|  | Hospital D | 11.6% | 17.6% | 27.6% | 14.0% | 28.3% |
| Non-COVID-19 | Hospital A | 3.0% | 17.9% | 9.0% | 4.0% | 26.8% |
|  | Hospital B | 2.7% | 11.8% | 17.2% | 4.4% | 34.4% |
|  | Hospital C | 3.6% | 13.0% | 21.4% | 7.7% | 33.0% |
|  | Hospital D | 3.1% | 6.9% | 11.5% | 2.0% | 23.1% |

Unsurprisingly, a substantially higher proportion of COVID-19 patients died and/or required ICU care and/or mechanical ventilation than non-COVID-19 patients.

### Raw Score AUC

The initial RI score calculated on admission for each patient was used to calculate the AUC for discrimination of patients with and without the target outcomes, independently evaluated, as shown in Table 5. For both COVID-19 positive patients and non-COVID-19 patients, the AUC values associated with the initial RI score range from good to excellent.

**Table 5.** AUC using initial RI score on admission to predict in-hospital mortality, ICU utilization, or need for mechanical ventilation.

| Population | Hospital | Mortality | ICU | Ventilation |
| --- | --- | --- | --- | --- |
| COVID-19 | Sinai LifeBridge | 0.79 | 0.79 | 0.75 |
|  | Yale New Haven | 0.87 | 0.67 | 0.75 |
|  | Bridgeport | 0.87 | 0.67 | 0.73 |
|  | Greenwich | 0.86 | 0.69 | 0.68 |
| Non-COVID-19 | Sinai LifeBridge | 0.88 | 0.85 | 0.89 |
|  | Yale New Haven | 0.90 | 0.84 | 0.88 |
|  | Bridgeport | 0.93 | 0.76 | 0.88 |
|  | Greenwich | 0.93 | 0.79 | 0.85 |

This performance is particularly notable in light of the fact that in many instances the target outcome may have occurred a considerable time (e.g. a number of days) following patient admission.

### RI vs. Age vs. Comorbidity for predicting inpatient mortality

To quantify the differences in the ability of the initial RI score to risk stratify COVID-19 patients compared with using either age or comorbidity, as measured by the Charlson Comorbidity Index (CCI), we computed the AUC of each measure for predicting in-hospital mortality, as shown in Table 6.

**Table 6.** AUC values for initial RI score on admission, patient age, and CCI (comorbidity) used as measures for predicting in-hospital mortality.

| Population | Hospital | RI | Age | CCI |
| --- | --- | --- | --- | --- |
| COVID-19 | Sinai LifeBridge | 0.79 | 0.68 | 0.67 |
|  | Yale New Haven | 0.87 | 0.78 | 0.70 |
|  | Bridgeport | 0.87 | 0.75 | 0.72 |
|  | Greenwich | 0.86 | 0.82 | 0.73 |
| Non-COVID-19 | Sinai LifeBridge | 0.88 | 0.68 | 0.71 |
|  | Yale New Haven | 0.90 | 0.73 | 0.74 |
|  | Bridgeport | 0.93 | 0.81 | 0.75 |
|  | Greenwich | 0.93 | 0.83 | 0.83 |

We note that the predictive performance of a single variable logistic regression model using the RI was not meaningfully improved by adding either age or CCI to the model.

### RI- Threshold-based performance

The precision and recall values of different RI threshold values were evaluated as shown in Figure 3. To orient the reader to Figure 3, the right-hand most points indicate a threshold of initial RI < 100, which encompasses all admitted patients, hence providing 100% sensitivity to mortality. Each point moving from right-to-left represents a 10-point reduction in RI threshold value. As expected, patients with lower RI scores, and hence higher acuity, on admission, have a greater probability of expiring in the hospital leading to an increasing precision at the expense of decreased recall.

**Figure 3.**
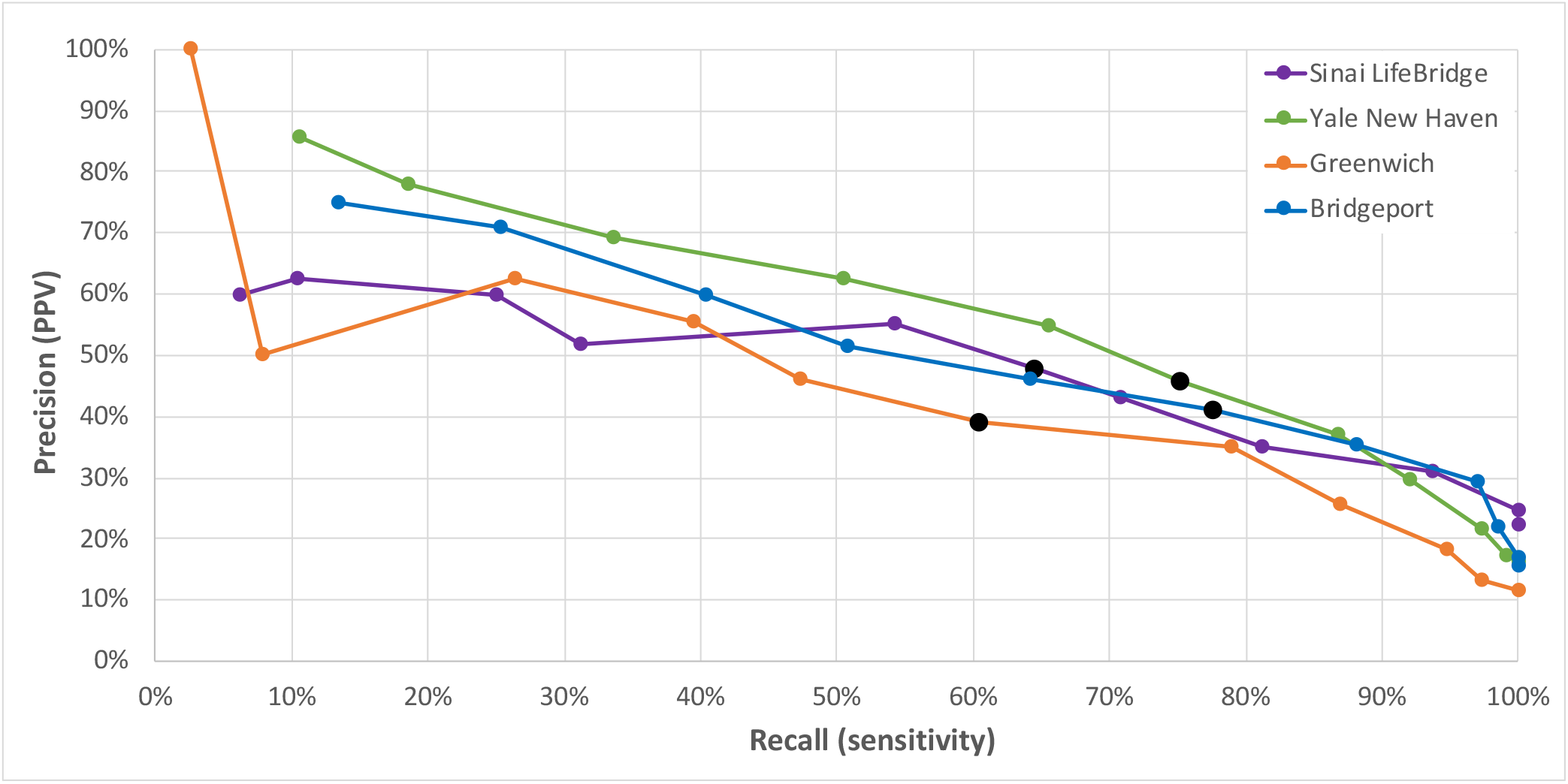
Precision-Recall for initial RI thresholds and in-patient mortality, in 10-point RI increments from 0 to 100 from left to right. Black points are a threshold value of RI< 50.

Operating point thresholds were selected to be initial RI < 50 to identify high risk patients at all facilities and initial RI > 70 (YNHHS hospitals) and initial RI > 75 (Sinai hospital) to identify low risk patients. RI scores from one organization to the next can vary due to differences in documentation practices, accounting for the slightly different low risk threshold values selected.

Table 7 (hospitals re-ordered and anonymized) details the percentage of the COVID-19 populations flagged due to being below and above these thresholds, respectively, for each hospital, as well as in-hospital mortality, ICU utilization, and mechanical ventilation rates within these groups.

**Table 7.** Percentage of COVID-19 patients and corresponding in-hospital mortality, ICU utilization, and mechanical ventilation rates stratified by high and low risk RI criteria.

| Hospital | HIGH RISK |  |  |  | LOW RISK |  |  |  |
| --- | --- | --- | --- | --- | --- | --- | --- | --- |
|  | RI < 50 (YNHHS), RI < 50 (Sinai) |  |  |  | RI > 70 (YNHHS), RI > 75 (Sinai) |  |  |  |
|  | COVID-19 patients | Mortality rate | ICU utilization rate | Mechanical ventilation rate | COVID-19 patients | Mortality rate | ICU utilization rate | Mechanical ventilation rate |
| Hospital A | 29.7% | 40.9% | 38.6% | 25.2% | 46.7% | 1.0% | 14.0% | 3.5% |
| Hospital B | 26.7% | 45.7% | 41.4% | 22.0% | 47.7% | 2.7% | 16.3% | 2.7% |
| Hospital C | 30.1% | 47.7% | 56.9% | 41.5% | 40.3% | 8.0% | 9.2% | 8.0% |
| Hospital D | 17.9% | 39.0% | 27.1% | 23.7% | 58.1% | 2.1% | 11.0% | 8.9% |

While fewer than a third of COVID-19 patients meet the high risk RI criteria on admission, they have a 39%-48% mortality rate, compared with approximately half of the COVID-19 population which meets the low risk RI criteria with a mortality rate of only 1-8%. Comparably large differences in ICU utilization rates and mechanical ventilation rates between the high and low risk groups are also evident.

Differences in the mean and median length of stay between the two risk groups is presented in Table 8. For three of the facilities we see that among patients who do not expire in the hospital, the low risk group has approximately half the length of stay as the high risk group (Greenwich hospital is the outlier in this regard, but as there were only four expired patients in the low risk group the results are not statistically meaningful). In contrast, for patients expiring in the hospital, length of stay is longer in the low risk group than the high risk group, implying that these low risk patients have a lower risk at the time of admission but suffer a critical deterioration event much later in the course of their hospitalization.

**Table 8.** Length of stay in days (mean | median) for all high and low risk COVID-19 patients and expired and non-expired sub-sets.

| Hospital | HIGH RISK |  |  | LOW RISK |  |  |
| --- | --- | --- | --- | --- | --- | --- |
|  | RI < 50 (YNHHS), RI < 50 (Sinai) |  |  | RI > 70 (YNHHS), RI > 75 (Sinai) |  |  |
|  | All high risk COVID-19 | Expired only | Non-expired only | All low risk COVID-19 | Expired only | Non-expired only |
| Sinai LifeBridge | 13.0 \| 10.0 | 9.8 \| 6.7 | 16.0 \| 13.4 | 6.4 \| 5.7 | 11.1 \| 9.2 | 6.0 \| 5.0 |
| Yale New Haven | 9.6 \| 7.7 | 7.3 \| 4.2 | 11.5 \| 10.2 | 7.7 \| 6.1 | 15.6 \| 11.2 | 7.5 \| 6.1 |
| Bridgeport | 8.4 \| 7.5 | 5.5 \| 4.3 | 10.4 \| 9.6 | 6.5 \| 5.1 | 9.5 \| 9.5 | 6.5 \| 5.0 |
| Greenwich | 7.5 \| 5.3 | 5.6 \| 5.1 | 8.7 \| 5.9 | 6.4 \| 5.1 | 7.8 \| 2.6 | 6.3 \| 5.1 |

### Opportunity to augment level of care decisions

To determine if COVID-19 patients deemed high risk based on initial RI were already recognized and treated as such by providers, we use patient location (i.e. ICU versus non-ICU) as a proxy for provider concern. Table 9 (reordered and anonymized) provides the ICU status of patients admitted in the high risk (initial RI < 50) category. We see that there are two potential opportunities for improvement: (a) direct admission or earlier transfer to the ICU for patients who ended up in the ICU, and (b) admission or transfer to the ICU for patients who never went to ICU but might have benefited from that level of care. We also see that for those high risk patients who expired in the hospital the time from admission to death ranged from a median of approximately four to seven days.

**Table 9.** Breakdown of ICU utilization and time to transfer (all ICU transfers) and time to death (all deaths) for COVID-19 patients meeting RI high risk criteria.

| Hospital | Direct Admit to ICU | Direct Admit to ICU who Expired | Transferred to ICU | Transferred to ICU who Expired | No time in ICU | No time in ICU who Expired | Days from admit to ICU Transfer<br>mean \| median | Days from admit to death (all deaths)<br>(mean \| median) |
| --- | --- | --- | --- | --- | --- | --- | --- | --- |
| Hospital A | 29.1% | 37.8% | 9.4% | 33.3% | 61.4% | 43.6% | 3.0 \| 1.5 | 5.5 \| 4.3 |
| Hospital B | 32.8% | 52.5% | 8.6% | 68.8% | 58.6% | 38.5% | 3.9 \| 2.4 | 7.3 \| 4.2 |
| Hospital C | 35.4% | 73.9% | 21.5% | 50.0% | 43.1% | 25.0% | 4.4 \| 2.9 | 9.8 \| 6.7 |
| Hospital D | 22.0% | 30.8% | 5.1% | 100.0% | 72.9% | 37.2% | 3.6 \| 3.9 | 5.6 \| 5.1 |

### Evaluation of 24-hour in-patient mortality

Giving critical consideration to the differences between RI at admission for COVID-19 and non-COVID-19 patients raises the question of whether the RI itself is insensitive to the particular physiological characteristics of COVID-19 patients. To test this, we assessed all RI scores during each patient’s admission to evaluate the RI as a predictor of 24-hour mortality for both COVID-19 and non-COVID-19 patients. Figure 4 shows almost no difference in the performance, which is expected as the physiologic deterioration preceding death, including vital sign derangement and organ system dysfunction, results in a concomitant increase in acuity that is largely independent of the underlying disease state.

**Figure 4.**
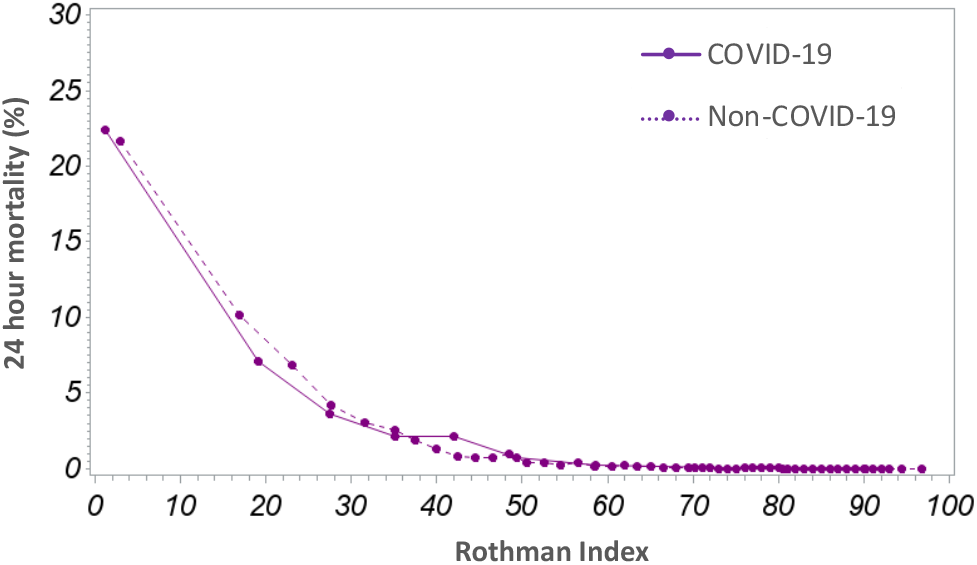
24-hour mortality vs RI for COVID-19 and non-COVID-19 patients at Sinai LifeBridge.

This confirms that the RI is correctly reflecting acuity for the COVID-19 population.

## Discussion

Initial reports of COVID-19-related mortality from China focused on the strong influence of patients’ age on acuity and mortality. These reports were followed quickly by data on the significance of co-morbidities in mortality rates. Yet other work has noted that while age appears to be a risk factor for COVID-19, it does not effectively discriminate between patients who are at highest risk or require intensive care unit level of support and those who do not.^2^ These observations match our finding that age is a poor prognosticator of the patients who will expire in the hospital. These observations can all be accounted for by recognizing that age is a correlate or crude proxy for comorbidities; comorbidities are a correlate and somewhat less crude proxy for acuity, but acuity bears a direct relation to mortality risk. Our work uses the Rothman Index to directly evaluate acuity in order to provide a more accurate and graduated assessment of mortality risk and by extension insight into the likely need for extreme supportive measures including mechanical ventilation or ICU level care.

The distinction in COVID-19 and non-COVID-19 curves for the RI on admission, seen in Figure 1(a), reflects the *high risk of subsequent critical deterioration* for COVID-19 patients relative to the general medical patient population, but we see that when the critical deterioration actually occurs, the manifestation of physiological derangement and decompensation in acuity appears the same as for any other patient in critical condition, as reflected by the close correspondence of the RI curves for predicting imminent (i.e. 24-hour) mortality in Figure 4. This lends credence to the idea that the RI correctly reflects COVID-19 acuity, but clinicians must re-calibrate their sense of what level of risk that acuity implies for COVID-19 patients at admission.

This work also quantifies and substantiates what has been anecdotally reported by front-line clinicians caring for COVID-19 patients, which is that after admission, these patients have substantially higher potential for significant deterioration than non-COVID-19 counterparts.

> *“Symptoms of COVID-19 may vary, and may include fever, chills, headache, myalgia, rhinorrhea, throat pain, dyspnea, chest pain, cough, and sputum. Less frequent symptoms are loss of smell and taste, nausea, vomiting, and diarrhea. The clinical course trends toward two different stages. The first typically includes these symptoms, and is primarily experienced during the first week after onset of symptoms. For some, the second stage starts between days five and seven when sudden rapid clinical deterioration may occur. We have not yet found any predictive symptoms of subsequent deterioration.”* ^30^

We see clear evidence of this in all four of the hospitals in the study when comparing the mortality rate, ICU utilization, and length of stay for COVID and non-COVID-19 patients admitted with similar initial acuity levels. This means that the clinician’s sense of how to treat a COVID-19 patient could be led astray by how they would treat a non-COVID-19 patient of similar acuity. Many COVID-19 patients who would not normally be viewed as “high risk” should be. This is the specific challenge of the “patients in the middle” – those who are not unequivocally of very low or very high acuity.

A further quantitative measure which supports our hypothesis that relevant acuity is not adequately captured by comorbidities is a comparison of the mean Charlson Comorbidity Index between COVID-19 patients who expire and non-COVID 19 patients who expire. For patients who expired in the hospital, the comorbidity burden at admission, as measured by the CCI, was less for COVID-19 patients than it was for non-COVID-19 patients. This is yet another measure which demonstrates that COVID-19 patients who expire don’t present with the same apparent degree of risk as non-COVID-19 patients who expire.

This work shows that the RI may be a factor which provides predictive utility regarding the risk of subsequent and sometimes unexpected deterioration in COVID-19 patients. Other ongoing research has also identified the initial RI as a meaningful indicator of COVID-19 patient risk.^31^ What we see from our analysis is that the RI can be used to clearly differentiate the high and low risk patients from among many of those patients “in the middle” where the prognosis of the patient is otherwise far from clear.

As this is a retrospective analysis it is important to understand the practical value of these findings. The obvious question is this: are the high risk patients already receiving appropriate and timely care? While we did not perform chart reviews, the fact that a high proportion of patients who met the RI high risk threshold and were only later transferred to, or indeed never admitted to, ICU, but nevertheless subsequently expired suggests that these placement decisions were not strictly made on the basis of patient acuity or risk. Decisions may have been the result of either a misreading of the severity of the patient’s condition or based on a determination that the patient would not benefit from ICU level care for either physiological reasons or given the patient’s or family’s wishes.

However, it is possible that some patients are not assigned to locations that can provide adequate attention and care. It is clear from the AUC values in Table 5 that acuity on admission is highly determinative of subsequent clinical deterioration. Specifically, 25-44% of those COVID-19 patients who met high risk RI criteria at admission subsequently expired but were never admitted to an ICU. Further we see in Table 9 that the median time from admission to expiration for high risk COVID-19 patients ranges from 4-7 days and for those high risk patients who were transferred to the ICU, the time from admission to ICU transfer ranged from a median of 1.5 to 4 days. This implies that if the physician was alerted to the risk implied by the initial RI, there may have been an opportunity to affect outcomes by altering care.

Perhaps equally important, the low risk group accounts for nearly half of all hospitalized COVID-19 patients but has a substantially lower rate of in-hospital mortality, ICU utilization, and mechanical ventilation than the high risk group. Additionally, from Table 8 we see that the vast majority of low risk patients do not expire and this group has approximately half the length of stay of the non-expiring high risk population. The opportunity to more critically assess the needs of the low risk patients relative to their high risk counterparts may help guide difficult decisions to transfer or discharge these patients into lower or non-acute care settings in order to free hospital and ICU bed capacity for use with higher risk patients.

Aligning level of care decisions at admission with hospital and ICU capacity constraints is a critical undertaking during COVID-19 surge conditions. Having reference to an objective measure may be helpful to ensure that patients are placed appropriately and resources are allocated efficiently. This applies equally to the high risk patients who may benefit from closer monitoring or more intensive therapies, and to lower risk COVID-19 patients, some of whom may not need high levels of care, or indeed hospitalization at all.

To that end, researchers, physicians, and nursing leadership at Sinai LifeBridge, the Yale-New Haven Health System, and other hospitals are exploring the use of the RI explicitly for risk stratification in the event of a further surge of COVID-19 patients.

### Limitations of the study

This work was a retrospective analysis of COVID-19 patients, and as such presupposes knowledge of whether or not a patient was accurately diagnosed with COVID-19. This work does not account for possible coding error nor uncertainty arising from the possibility of false positive or false negative diagnostic lab tests. However, with the dramatic improvement in testing capacity it is reasonable to expect that diagnoses are correct in most instances. Additionally, the care of COVID-19 patients is evolving rapidly, and treatments such as dexamethasone, remdesivir and convalescent plasma, proning, and anti-coagulation therapies are now being used in a manner that was not routine during the period of our study data^32-38^ while the benefits of early, aggressive intubation are increasingly debated.^39,40^ Without chart review, it is not possible to know what patient goals of care may have been and the extent to which they influenced decisions related to treatment or to receiving care in the ICU. We also note that the data reflecting non-COVID-19 cases largely overlaps with a time period when most U.S. hospitals were taking extraordinary measures to manage population volumes and hence the medical admissions during this time will not be perfectly representative of typical non-COVID-19 medical patient populations.

### Conclusion

COVID-19 patients have high mortality rates compared to other medical service patients. They are subject to rapid deterioration which may follow days after hospital admission. Providing clinicians with early predictive insights into which of these patients are most at risk for such deterioration would usefully support admission and care decisions. A study of 1,669 COVID-19 patients at four hospitals has shown that the Rothman Index, a patient acuity score, can be used at the time of admission to stratify risk of critical deterioration in COVID-19 patients. This measure may be an important supplement to physician judgement, as non-COVID-19 and COVID-19 patients who present with similar acuity may not have the same risk for poor outcomes, and hence, the same requirements for care. As the United States prepares for prolonged stress from the COVID-19 pandemic, there are significant implications to being able to rapidly and objectively support clinicians in their efforts to determine COVID-19 patient risk and appropriate care settings at the time and point of patient admission.

## Data Availability

This work entailed an IRB approved study of retrospective data.

## Acknowledgements

We extend our thanks to Christine Sullivan and Lynn Lewis of Sinai LifeBridge Hospital for their help to make this work both feasible and useful, and to Dr. Rob Fogerty of Yale New Haven Hospital for his thoughtful insights and suggestions.

